# Online dashboards for SARS-CoV-2 wastewater data need standard best practices: an environmental health communication agenda

**DOI:** 10.1101/2022.06.08.22276124

**Authors:** Colleen C. Naughton, Rochelle H. Holm, Brooklyn P. James, Ted Smith

**Affiliations:** Department of Civil And Environmental Engineering, University of California Merced, 5200 Lake Rd., Merced, CA 95343, United States; Christina Lee Brown Envirome Institute, School of Medicine, University of Louisville, 302 E. Muhammad Ali Blvd., Louisville, KY 40202, United States

**Keywords:** COVID-19, health communication, sewage, standards, technology, wastewater-based epidemiology

## Abstract

The rapid development of scientific communication approaches for environmental surveillance data with online information dashboards has been done in the absence of a global organizing body during the coronavirus disease 2019 pandemic. We aim to make a case for standardization of dashboards presenting SARS-CoV-2 wastewater data. The list of dashboards was compiled as of March 31, 2022. The 127 dashboards reviewed represented 27 countries using a range of line/bar graphs, maps, and tables with symbol presentation. We identified 96 separate units of measure for the wastewater SARS-CoV-2 data. There was also inconsistency in using linear or log scale. Twenty-five percent of dashboards presented SARS-CoV-2 variant monitoring. Only 30% (38/125) of dashboards provided downloadable source data. There is great opportunity to improve scientific communication though the adoption of uniform data presentation conventions or standards for this field.

## 1. Introduction

Wastewater surveillance for severe acute respiratory syndrome coronavirus 2 (SARS-CoV-2) has been used since early in the coronavirus disease 2019 (COVID-19) pandemic to complement clinical testing.^1–3^ Online data dashboards are a one-way message from experts to both experts and nonexperts. Surveys of public awareness and perceptions of wastewater surveillance have reported strong support for wastewater data sharing.^4 5^ The current context for our research agenda is that risk communication problems may come from establishing and recognizing credibility, making messages understandable, rapidly preparing messages, capturing, and focusing attention, and getting data.^6^ Stakeholders including the World Health Organization^3 7^ have promoted online information dashboards for presenting SARS-CoV-2 wastewater viral concentration. Yet, the historical context of getting risk communication right for scientists has been evidenced when failing to evaluate and communicate to the public earthquake hazard data resulted in scientist and government officials going on trial for the 2009 L’Aquila, Italy, earthquake.^8^ The rapid development of scientific communication approaches for environmental surveillance data with online information dashboards has been done in the absence of a global organizing body during the COVID-19 pandemic despite efforts convene the field to address this issue among others.^9^

We reviewed 127 presentation formats of SARS-CoV-2 wastewater data on global online dashboards for communicating public health risk. Additionally, we critically reflect on the gaps in the underlying science to advocate for an improved environmental health communication agenda.

## 2. Surveying data landscape

Data were collected from the COVIDPoops19 global dashboard of wastewater monitoring for SARS-CoV-2^10 11^ as of March 31, 2022. For non-English dashboards, Google translate was used. This study was conducted prior to the United States Centers for Disease Control and Prevention^12^ National Wastewater Surveillance System public dashboard. To determine country level income classifications, the World Bank^13^ guidelines were used. Dashboards used in the analysis are available online public records, links of which are provided in Supplement A. Data provided in Figure 1 is covered under the University of Louisville Institutional Review Board is classified as non-human subject research (reference #: 717950).

**Figure 1.**
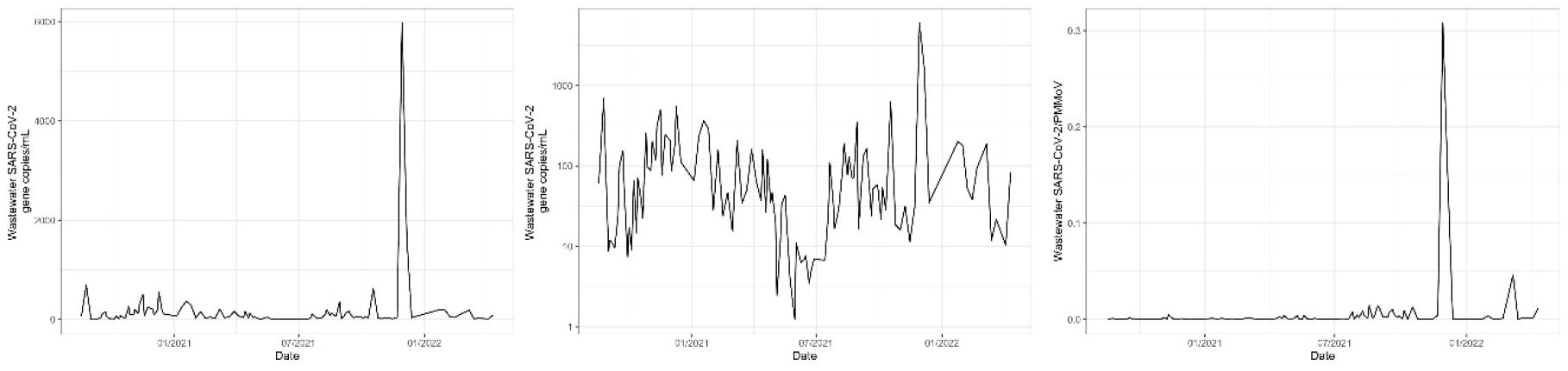
Three time-series plots showing SARS-CoV-2 viral wastewater, August 2020 to April 2022: Left panel displays wastewater data on a linear scale; Middle panel displays wastewater data on a logarithmic scale; and Right panel displays wastewater data normalized by PMMoV.

## 3. Current data presentation

The focus of this research agenda is advocacy for improved scientific communication in online dashboards presenting SARS-CoV-2 viral concentration in wastewater. From 127 dashboards identified, 125 were accessible at the time of this analysis (two websites previously tracked had been moved or were offline). More than half (58%; 72/125) provided SARS-CoV-2 wastewater and COVID-19 case data on the same dashboard.

### 3.1 Dashboard locations

Twenty-seven countries had COVID-19 wastewater dashboards, 94% (118/125) of which were high-income countries. While some high-income countries had a single dashboard, others had many; within the United States there are 66 unique dashboards alone, Canada has 22 dashboards, and Australia had five dashboards. The remaining dashboards (6%; 7/125) belonged to low- and middle-income countries, and still Brazil and South Africa had two unique dashboards, respectively. Multiple dashboards within a country using different metrics suggests an opportunity for standards.

### 3.2 Units of measure

Data was presented across a range of formats including line/bar graphs, maps, and tables with symbols. We identified 96 separate ways SARS-CoV-2 wastewater units of measure were written (Table 1). Of these, some form of quantitative (in contrast to presence/absence or trend observation) wastewater units results were given in 85% (106/125) dashboards. The more common was a derivative of gene copies/L or gene copies/mL (27%; 34/125). Differences in units can be confusing. The <u>Erie County, PA</u>, dashboard reported “COVID-19 Amounts in Wastewater” which confuses the resulting disease with the indicator cause.

**Table 1.** Select SARS-CoV-2 wastewater concentration unit groups as presented in online dashboards.

| Group | Example units |
| --- | --- |
| Viral concentration | cp/mL<br>COVID-19 amounts in wastewater<br>genome copies/L<br>genome copies/L of sewage<br>genome copies/mL wastewater<br>genome per Liter<br>log10 gene copies/L<br>log10 RNA Copies/L<br>mean virus copies/L<br>relative SARS-CoV-2 levels<br>SARS-CoV-2 RNA copies/mL<br>SARS-CoV-2 (weighted mean; gc/L)<br>standardized concentration of SARS-CoV-2 Gene Copies<br>RNA copies/mL<br>virus copies/L<br>wastewater viral load (Log10 Copies) |
| Viral concentration normalized by population size sampled | # virus particles/100,000 inhabitants<br>copies SARS-CoV-2 per 100K<br>gene copies/day/100,000 people<br>log10 Copies per day per person<br>M copies/person/day<br>SARS-CoV-2 RNA copies/day/100 000 eq. inhabitants<br>µg/L per 100 thousand inhabitants<br>viral gene copies/person |
| Viral concentration normalized by time | average covid million genes per day<br>EWMA copies per day<br>gc/day<br>N1, N2, IP4 GC/Day<br>SARS-CoV-2 RNA flux (copies/day) |
| Viral concentration normalized by a fecal indicator positive control | concentration of SARS-CoV-2/PMMoV<br>copies/mL normalized to PMMoV copies/mL x 1000<br>copies/PMMoV<br>N1 gene copy number to PMMoV ($\times 10^4$)<br>normalized viral copies (to PMMoV)<br>normalized viral copies ($\mu$)<br>PMMoV normalized N1 ($\times 1000$)<br>ratio of gene N1 and PMMoV |

Manipulation of the y-axis for presentation was also common. Population adjustment was done by the number of people served by the sewer service area sampled (for example, by 100,000 inhabitants) (15%; 15/102), less frequently by the positive control fecal indicator pepper mild mottle virus (PMMoV) (8%; 8/102), or by sewer system flow rate (2%; 2/102). Some dashboards, such as <u>Davis, CA</u>, dashboard had a non-numeric y-axis attributed to the normalization ratio. Two other dashboards displayed data in the form of exponentially weighted moving average (EWMA) per day (<u>Bengaluru (Bangalore), India</u> and <u>Missouri</u>), which again may be difficult in data interpretation for the public. More pertinently is the absence of a y-axis unit, such as the dashboard by the <u>PA Department of Corrections</u>.

### 3.3 Scaling

Where units were presented (Figure 1), a linear scale only was observed more often (63%; 64/102) than a logarithmic scale (27%; 28/102). Ten dashboards (10%; 10/102) presented both logarithmic and linear scales. Some (34%; 35/102) dashboards with line/bar graphs had a double y-axis, commonly with COVID-19 clinical, hospitalization, vaccine rate, or number of deaths thoughtfully scaled with wastewater data. However, wastewater concentration on the left y-axis and normalized data on the right y-axis was also present. Remaining dashboards had a non-numeric scale data presentation or used a trend indicator table with symbols.

Date was the most common scale of intersect, with sixty dashboards presenting data back to 2020, thirty-six back to 2021 and six starting in 2022. Not all dashboards use line/bar graph. For example, the New South Wales, Australia, dashboard uses symbols to denote stable, increasing, and decreasing in a table format for sample location by date while the Canadian Water Network Wastewater Collection Maps uniquely presented a partnership dashboard on sample location lab processing samples (public lab or university lab), though with no wastewater data.

### 3.4 Variant monitoring

One quarter (25%; 31/125) of dashboards presented data on SARS-CoV-2 variant tracking, mostly (94%; 29/31) were from high-income countries. Variant monitoring was more typically displayed in a separate dashboard section, by comparing the percentage of each variant type. The variant tracking dashboard presented ranged for example from Switzerland using digital droplet polymerase chain reaction assays to present derived estimates of variants, to Jefferson County, KY, presenting results from genomic sequencing.

### 3.5 Data transparency

Although online dashboards could be viewed as a form of scientific communication, only 30% (38/125) provided a downloadable source file. This ranged from quantifiable data as a Microsoft Excel file in a variety of row and column formats to figures in a portable network graphic file format. In general, data transparency was observed to be low across the dashboards surveyed.

## 4. Discussion

The current landscape of data presentation for SARS-CoV-2 wastewater data through online dashboards found a lot of variation which could be addressed through standards. Environmental data dashboards have been shown to be a positive method for engaging stakeholders and supporting water and electricity sustainability methods, and a way to bridge between academics to communities being surveilled.^14^ For clinical samples, the real time dashboard by Johns Hopkins University is an example of individual diagnostic testing results aggregated from many global sources with a unified presentation;^15^ but the principles for rapid and responsible data sharing in epidemics have been focused on clinical testing results^16^ rather than wastewater analogs. Additionally, there remain more geographic areas where wastewater data collection is happening without an online dashboard.^11^ If there is variability across dashboards within a country in the way the data are calculated or presented, it could hamper understanding of transmission dynamics within and across borders. A member of the public traveling for work or vacation wanting to know about community spread can easily look at clinical case data across geographic locations for comparable results, whereas looking across wastewater dashboards for another location is challenging and inconsistent. We found few common dashboard themes, take-aways, and how public health principles are woven throughout each. Even the different impressions of scale should be emphasized,^17 18^ where the doubling of values in a linear scale is muted on a log scale. This contrasts with other environmental surveillance data such as *Escherichia coli* in drinking water whereby the World Health Organization guideline is 0 colony-forming units (cfu) per 100 mL,^19^ and the y-axis would most commonly be in cfu/100 ml but the x-axis could routinely be date or sample location. In microbiology, log scales are the norm attributed to a dynamic range of multiplying organisms and being above a drinking water guideline value is most important, whereas for SARS-CoV-2 wastewater trends over time is important. Changing between a log and linear scale of a progressively dynamic range of COVID 19 metrics has been shown to alter the public’s perceptions and policy preferences.^17 18^

We found 86% of the SARS-CoV-2 dashboards studied provided a quantitative wastewater result, this contrasts with risk communication around polio detection in wastewater, where presence/absence is the level of detail required.^20^ When presented in line/charts, the predominate y-axis was a derivative of gene copies/L or gene copies/mL against an x-axis of date. Flexibility should be available for normalization due to industrial flow, but normalization should be clearly stated, and non-transformed data should still be available.

The extent of the data presentation problem may not be fully understood at present, reflecting on poor hazard data communication from scientist and government officials from the L’Aquila, Italy, earthquake^8^ gives both insight and motivation to get this right. While SARS-CoV-2 viral concentration could be considered the basic level of data transparency, variant monitoring has evolved over the pandemic frameworks.^21^ Variant monitoring requires higher level laboratory facilities which may not be available in all public health centers, and standardization may be more nuanced.

The broader science community is better served to promote accuracy and reduce bias when there is a capability of transforming source data as a post hoc means of arriving at comparable data, so our advocacy for downloadable machine-readable data should be valuable to support that mission. Provision of wastewater source data should also adhere to (Findability, Accessibility, Interoperability, and Reusability) open data standards, and many countries have signed open data agreements.^11^ Data presentation does not solely need to be presence of a viral concentration, there is also excellent value in indicating the absence (or a quantified result below detection). This is an area requiring thoughtful global standardization.

Key considerations for a new standard format for online dashboards presenting SARS-CoV-2 wastewater data:

- Standards should apply globally
- Units of measure: labeled y-axis as log gene copies/L or gene copies/mL and labeled x-axis as date. If normalization is used, add a toggle to source data
- Machine-readable downloadable quantified source data
- It is never too late to start; researchers should be continually encouraged to add online dashboards

## 5. In Conclusion

We urge those at the forefront of dashboards for SARS-CoV-2 wastewater concentrations to standardize. There is an important need for clear and consistent communication for the non-research audience and specifically the greater public health community. The clearest example from our review of the need for standardization in the current landscape is the ninety-six different units from 125 dashboards and no clear precedent to follow; this is a significant issue and opportunity. But, if the field is trying for data transparency, not only have we not made it easy for non-experts, but more importantly, it is showing the underlying science still does not have a rationale for how we should calculate and present viral concentration. There also remains potential work on co-creation with community, utilities, and public health department input. These standards can be useful for other wastewater-based epidemiology applications as the field continues to expand beyond COVID-19. Our ability to incorporate wastewater into public health decision making is potentially hampered by the inconsistencies in the way data are analyzed and presented, but will we ever have one uniform standard?

## Data Availability

Dashboards used in the analysis are available online public records, links of which are provided in Supplement A.

## Contributors

Conceptualization: CCN, RHH, TS; Methodology: CCN, RHH, BPJ, TS; Formal analysis: RHH and BPJ; Writing-original draft preparation: RHH; Writing-review and editing: CCN, RHH, BPJ, TS; Supervision: TS; Project administration: TS. All authors have read and agreed to the published version of the manuscript.

## Patient consent for publication

Not applicable.

## Ethics approval

This study does not involve human participants.

## Supplement A

List of online dashboards for SARS-COV-2 wastewater from COVIDPoops19 website as of March 31, 2022

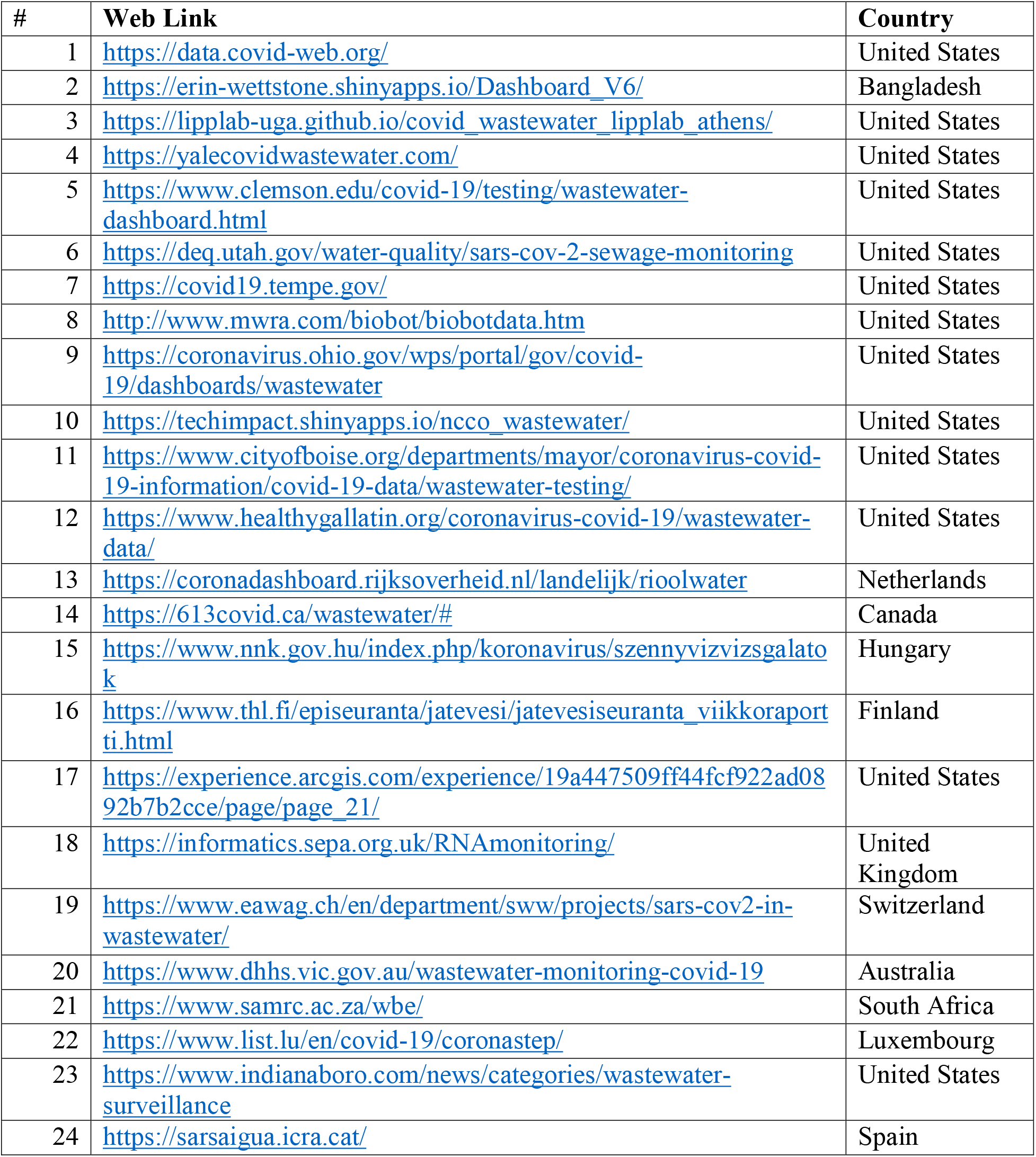
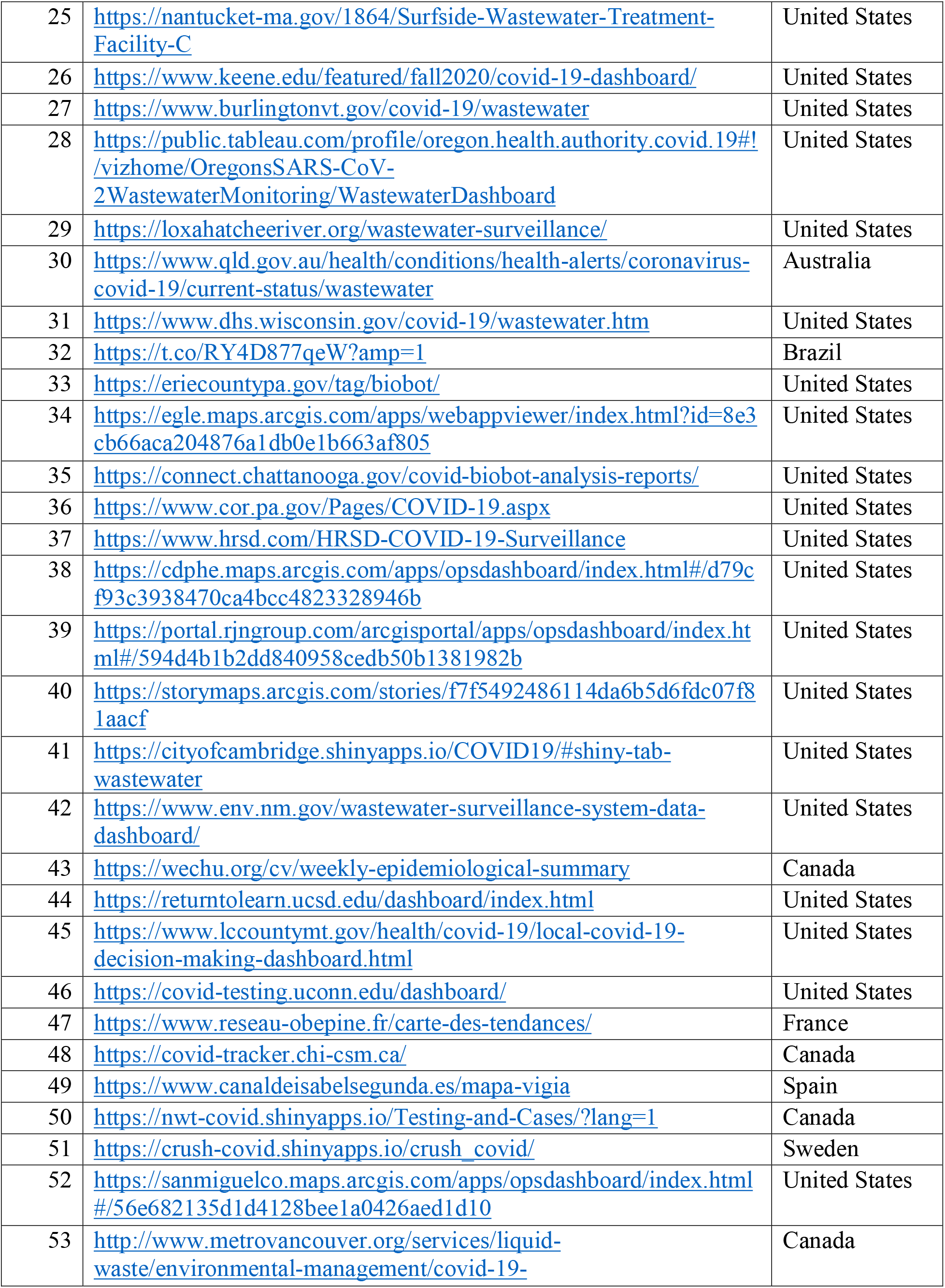
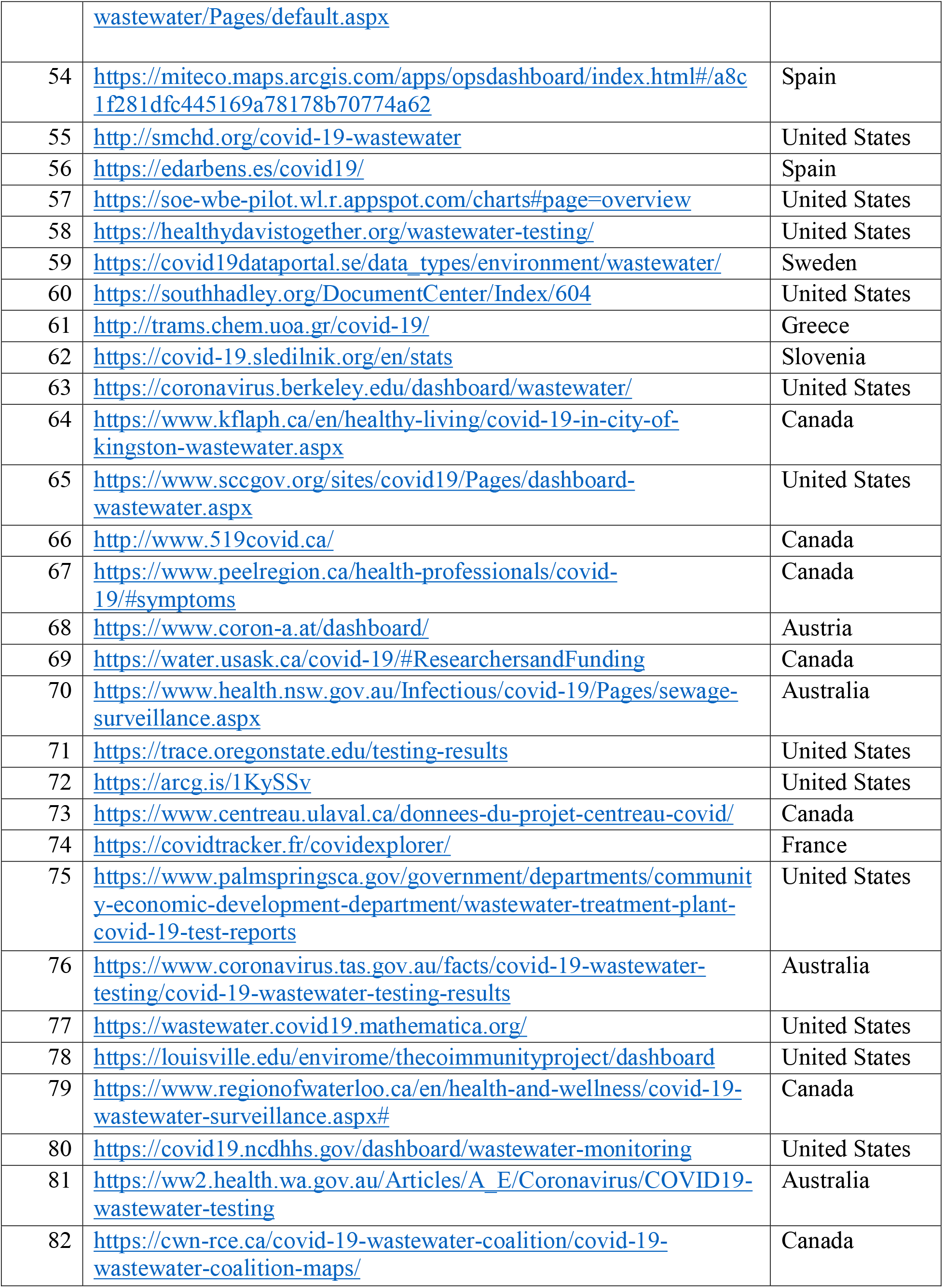
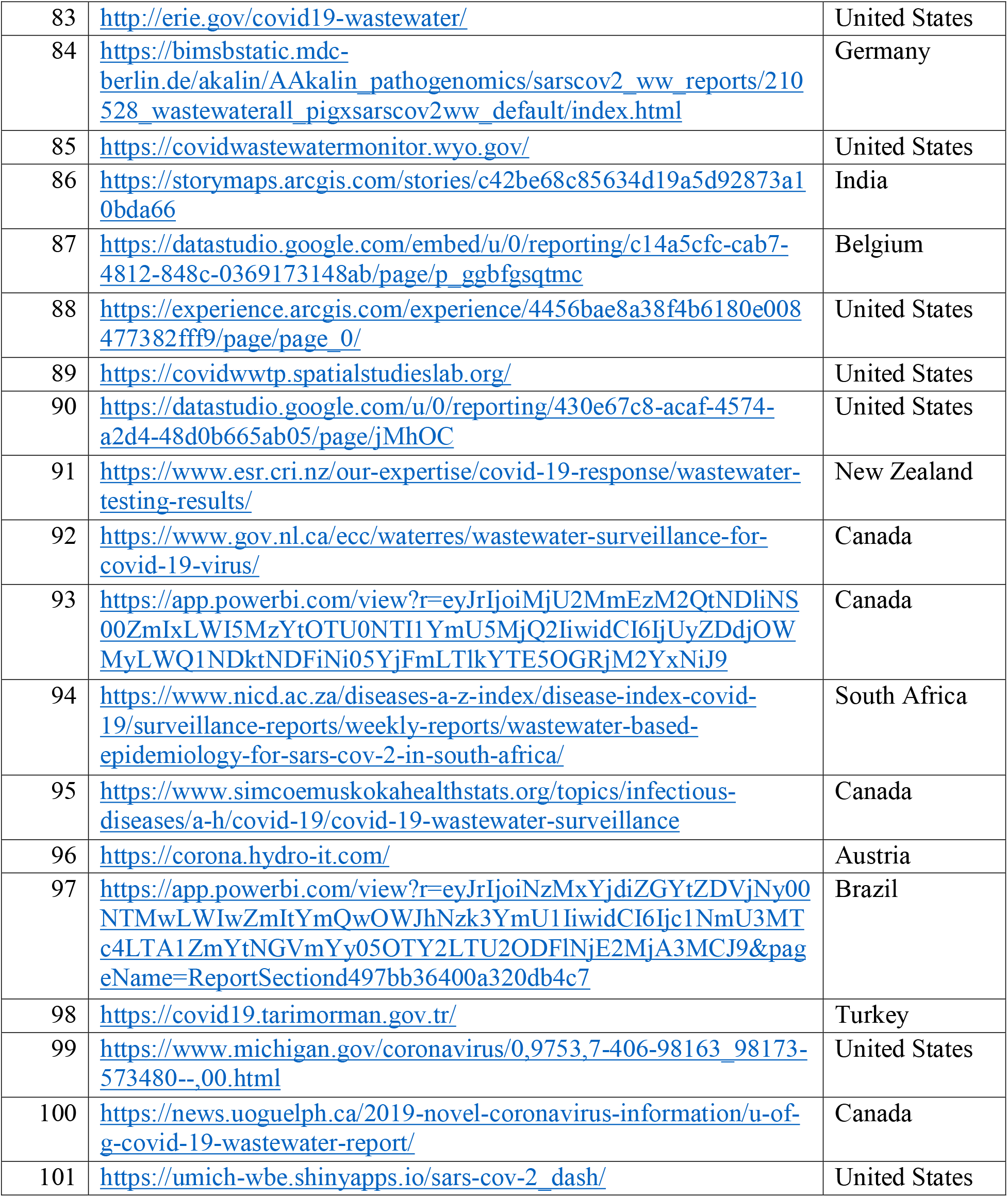
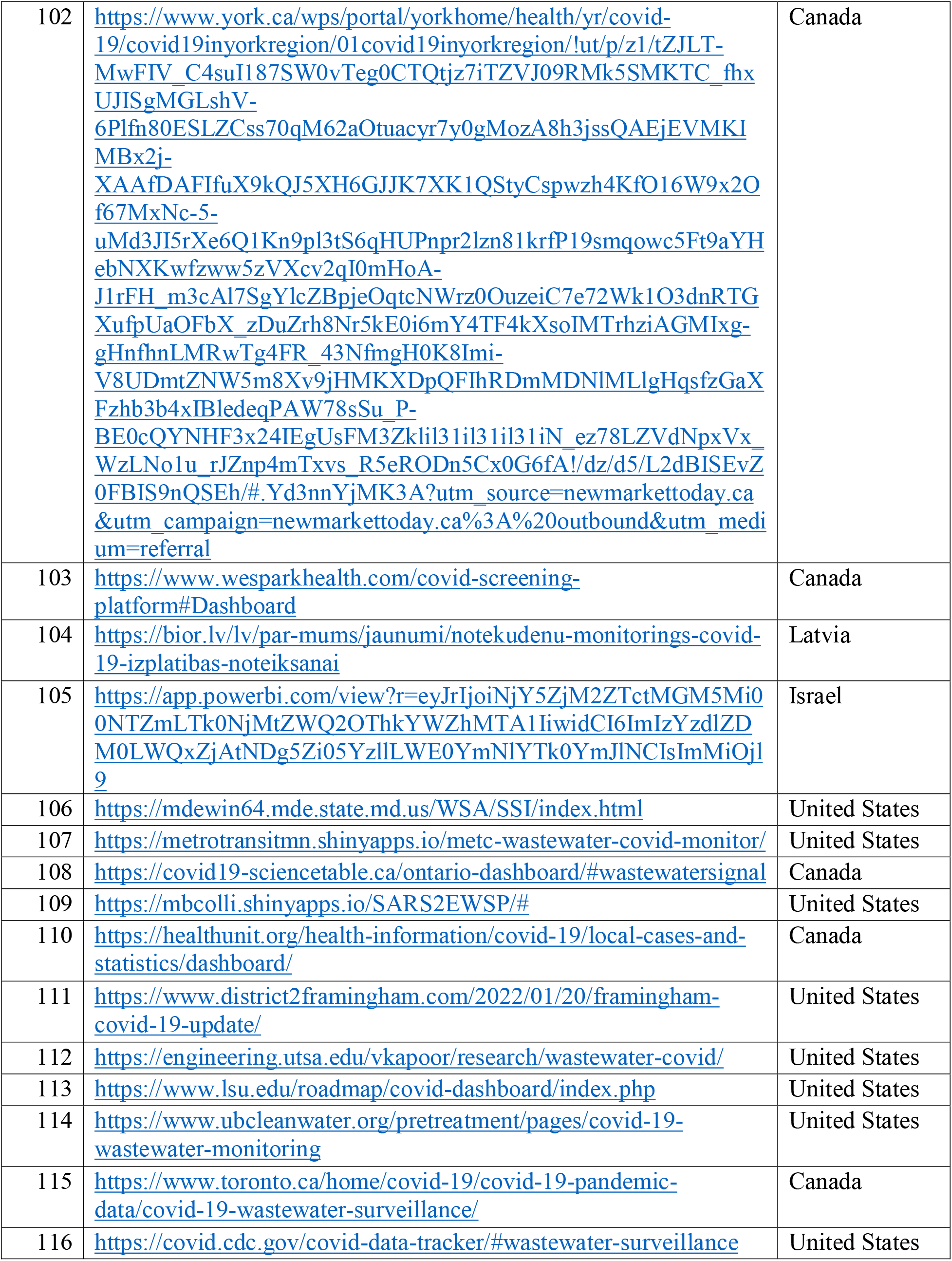
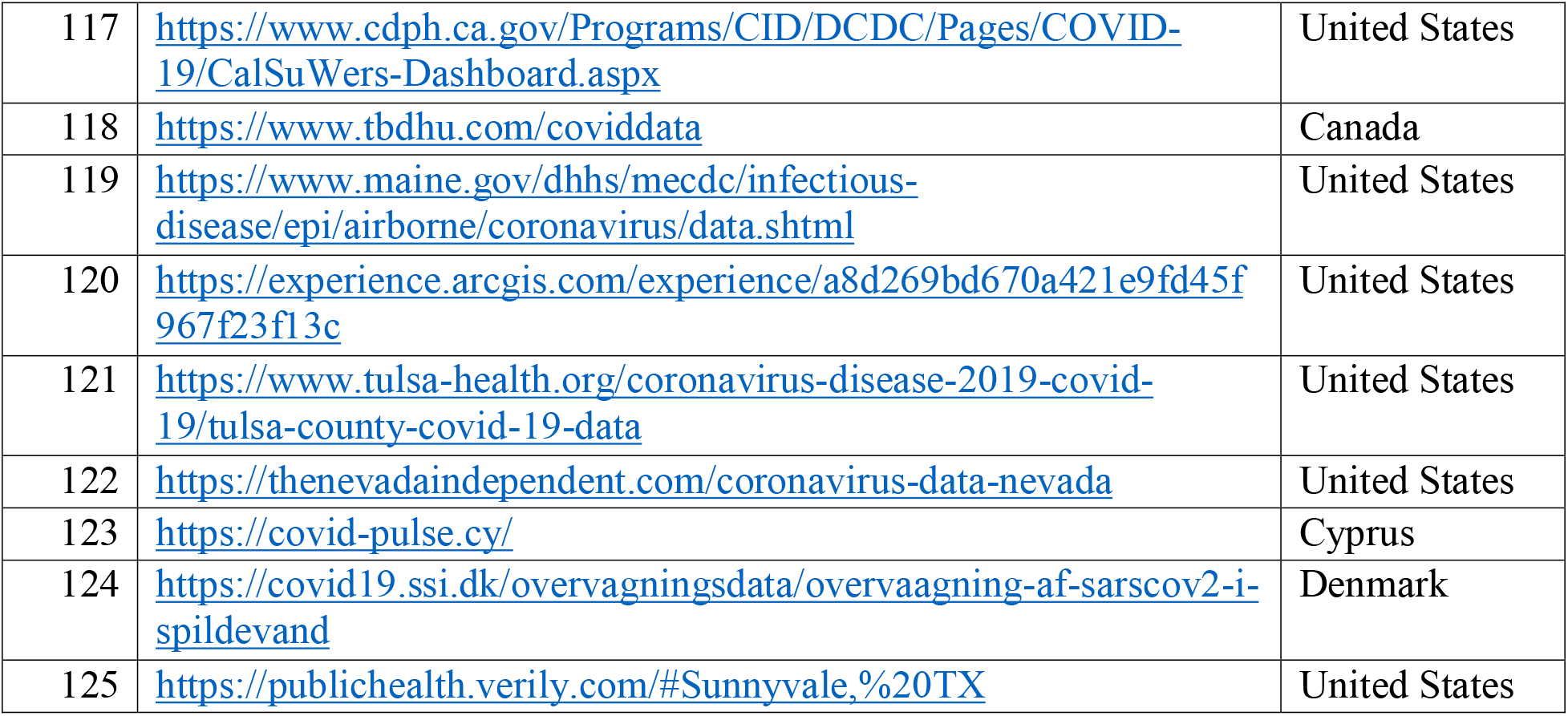

